# Resting-state functional connectivity alterations in obsessive-compulsive disorder: relationships between connectivity and clinical profiles in the Global OCD study

**DOI:** 10.1101/2025.07.30.25332428

**Authors:** Anders Lillevik Thorsen, Niels T. de Joode, Petra J.W. Pouwels, Céline N. Dietz, Feng Liu, Maria C.G. Otaduy, Bruno Pastorello, Frances C. Robertson, Jonathan Ipser, Seonjoo Lee, Dianne M. Hezel, Marcelo C. Batistuzzo, Marcelo Q. Hoexter, Marco A. N. Echevarria, Karthik Sheshachala, Janardhanan C. Narayanaswamy, Ganesan Venkatasubramanian, Christine Lochner, Euripedes C. Miguel, Y.C. Janardhan Reddy, Roseli G. Shavitt, Dan J. Stein, Melanie Wall, Odile A. van den Heuvel, Helen Blair Simpson, Chris Vriend

**Affiliations:** Bergen Center for Brain Plasticity, Haukeland University Hospital, Bergen, Norway; Center for Crisis Psychology, University of Bergen, Bergen, Norway; Amsterdam UMC, Vrije Universiteit Amsterdam, Department of Psychiatry, and Department of Anatomy and Neuroscience, de Boelelaan 1117, Amsterdam, the Netherlands; Compulsivity, Impulsivity and Attention, Amsterdam Neuroscience, Amsterdam, the Netherlands; Brain Imaging, Amsterdam Neuroscience, Amsterdam, the Netherlands; Amsterdam UMC, Vrije Universiteit Amsterdam, Department of Radiology and Nuclear Medicine, Amsterdam Neuroscience, de Boelelaan 1117, Amsterdam, the Netherlands; College of Life Sciences, University of Amsterdam, Amsterdam, the Netherlands; Columbia University Irving Medical Center, Columbia University, New York, NY 10032; The New York State Psychiatric Institute, New York, NY 10032, U.S.A; LIM44, Hospital das Clinicas HCFMUSP, Instituto e Departamento de Radiologia da Faculdade de Medicina, Universidade de São Paulo, SP, Brazil; Cape Universities Body Imaging Centre, University of Cape Town, South Africa; SAMRC Unit on Risk & Resilience in Mental Disorders, Department of Psychiatry & Neuroscience Institute, University of Cape Town, South Africa; Obsessive-Compulsive Spectrum Disorders Program, LIM23, Hospital das Clinicas HCFMUSP, Instituto & Departamento de Psiquiatria da Faculdade de Medicina, Universidade de São Paulo, SP, Brazil; Department of Methods and Techniques in Psychology, Pontifical Catholic University, São Paulo, SP, Brazil; National Institute of Mental Health & Neurosciences (NIMHANS), Bangalore, India; Institute for Mental and Physical Health and Clinical Translation (IMPACT), Deakin University, VIC, Australia; SAMRC Unit on Risk & Resilience in Mental Disorders, Department of Psychiatry, Stellenbosch University, South Africa

## Abstract

**Background:** Obsessive-compulsive disorder (OCD) has been associated with altered resting-state functional connectivity (FC), but findings are inconsistent and associations with clinical characteristics are unclear. The Global OCD study collected harmonized imaging, demographic, and clinical data across five sites and continents, with the aim of identifying consistent OCD-related alterations in FC.

**Methods:** We included 263 unmedicated adults with OCD and 254 healthy controls (HC) and estimated FC from resting-state functional magnetic resonance imaging. Between-group differences in and associations of clinical characteristics with FC were analyzed through general linear models adjusted for sex, age and years of education. We investigated seed-based FC of the amygdala, posterior putamen, nucleus accumbens, ventral and dorsal caudate nucleus, FC between and within atlas-based subnetworks, global network measures (global efficiency, modularity), and performed network-based statistics (NBS).

**Results:** Seed-based analysis showed weaker FC between the nucleus accumbens and left cerebellum in OCD versus HC, while NBS revealed a network of widespread hypoconnectivity spanning 249 regions. In OCD, FC was associated with previous use of selective serotonin/norepinephrine reuptake inhibitors, comorbid major depressive disorder, severity of anxiety, and sexual/religious and contamination symptoms. In particular, anxiety symptoms were linked to widespread stronger FC while sexual/religious symptoms were linked to stronger sensorimotor FC.

**Conclusions:** Individuals with OCD show widespread hypoconnectivity compared to HC, with considerable interindividual variation within OCD related to previous medication use, comorbid depression, severity of anxiety, and specific OCD symptoms. General hypoconnectivity in OCD may be a good target for interventions modulating brain circuit connectivity, including transcranial magnetic stimulation.

## Introduction

Obsessive-compulsive disorder (OCD) is characterized by intrusive thoughts, images or urges (obsessions) and/or repetitive mental or physical rituals (compulsions) and has a 1-3% lifetime prevalence rate, leading to considerable distress or functional impairment (1). OCD has been related to altered brain activation, structure, and connectivity of cortico-striato-thalamo-cortico (CSTC), frontolimbic and frontoparietal circuits (1–3). However, the alterations are inconsistent across studies and likely influenced by low statistical power, variation in data acquisition, processing, and analysis, and interindividual variation in symptom profiles. These shortcomings limit our ability to identify robust brain signatures and use neuroimaging to diagnose and treat OCD (1–3).

Resting-state functional magnetic resonance imaging (rs-fMRI) can be used to investigate how the brain functionally connects and communicates at different levels of organization (4, 5). There are several approaches to investigate functional connectivity (FC): 1) connectivity of seed regions with the rest of the brain, 2) connectivity between regions of interest or subnetworks based on atlas regions, or 3) use of graph theoretical models to estimate the topology of the functional “connectome” (6, 7). Seed-based approaches in OCD have found both stronger and weaker FC within and between different CSTC circuit regions in individuals with OCD compared to healthy controls (HC) (8–12). An atlas-based approach was recently used in a mega-analysis of 1024 adults with OCD and 1028 HC from the ENIGMA-OCD working group (13), and revealed widespread hypoconnectivity across the brain, as well as stronger connectivity between the thalamus, sensorimotor, visual, and prefrontal regions in individuals with OCD vs HC (13). At the connectome level, individuals with OCD have shown an imbalance between functional integration and segregation, as indicated by lower small-worldness, efficiency, and modularity (14–17). While there is evidence from smaller single-site studies that clinical interindividual variation is related to FC, including symptom severity (18), medication use (18, 19), symptom dimensions (20), insight (21), age of onset (22), and comorbid disorders (23), well-powered studies combining data from multiple sites (e.g. ENIGMA-OCD) are limited by their retrospective design and have not allowed nuanced investigation of this issue.

The Global OCD prospective study recruited a large sample of 281 medication-free adults with OCD and 265 HC matched on age and sex across five global sites on different continents (approximately 50 individuals with OCD and 50 HC at each site) (24). The study used extensively harmonized procedures for collecting and processing clinical, neurocognitive and MRI data, allowing for rich clinical phenotyping and careful standardization and maintenance of imaging parameters throughout the study and across sites (24–27). Recent results from the Global OCD study of structural connectivity and gray matter morphology suggest that case-control differences are small while highlighting the importance of clinical variables. For example, we found evidence for less fractional anisotropy in the posterior thalamic radiation and superior longitudinal fasciculus as well as lower global efficiency of the structural connectome in late-compared to early-onset OCD (28). Likewise, using FreeSurfer and voxel-based morphometry, we found few case-control differences in gray matter morphology but relevant associations between clinical variables (such as OCD severity and the presence of comorbid disorders) and morphometry of discrete cortical and subcortical regions (29).

The present study investigated FC using rs-fMRI in adults with OCD and HC, and its association with clinical characteristics within those with OCD. We used several approaches to investigate FC at the circuit, subnetwork, and connectome levels of brain organization using both pre-registered hypotheses and exploratory analyses (https://osf.io/b3vz5). Using seed-based FC, we first examined FC in the ventral cognitive, dorsal cognitive, sensorimotor, frontolimbic, and affective CSTCs. Here, we expected that adults with OCD, compared to HC, would show weaker FC between the dorsal cognitive (11, 20) and sensorimotor circuits (10, 11, 13). We also expected to find stronger FC between the ventral cognitive (20), affective (8, 11), sensorimotor (2), and frontolimbic circuits (8) in individuals with OCD compared to HC. Within the OCD group, we also expected to find weaker FC within the affective and frontolimbic circuits in those with an early (<18 years) versus late (≥18 years) onset of OCD (22, 30). Second, we used an atlas-based approach to compare FC between and within the seven subnetworks defined by Yeo et al. (31) expecting to find weaker connectivity within the somatomotor subnetwork in OCD versus HC (13). Within the OCD group, we also expected to find weaker connectivity within the limbic subnetwork in the early-versus late-onset of OCD (22, 30).

Exploratory analyses extended the seed-based and subnetwork-based analyses to all five CSTCs and seven Yeo subnetworks, respectively. We also compared OCD and HC and examined associations with clinical variables within OCD across the connectome using two approaches. We first used the Network-Based Statistics (NBS) approach, which seeks to identify clusters of connections that are associated with the independent variable (6). Secondly, we used graph theoretical measures to estimate network properties, where global efficiency quantifies information integration across the connectome while modularity reflects the presence of densely interconnected modules (7).

## Materials and methods

### Participants

The Global OCD study recruited 281 unmedicated adults with OCD and 265 HC aged 18-50 years old from five research sites in Brazil, India, the Netherlands, South Africa, and the USA (24). OCD was diagnosed using the Structured Clinical Interview for DSM-5 (SCID) (32) and obsessive-compulsive symptoms were measured using the Yale-Brown Obsessive-Compulsive Scale (Y-BOCS, minimum score 16) (33). Exclusion criteria for OCD participants included: psychotropic medication use (other than prn sleep medicines) or exposure and response prevention therapy in the previous six weeks; history of a psychotic/bipolar/anorexia nervosa/ Tourette disorders; or current substance use disorder, bulimia, or chronic tic disorder. Exclusion criteria for HCs included: a lifetime psychiatric disorder other than major depressive disorder or an anxiety disorder (but only if not in past year); history of psychotropic medications (other than prn sleep medicines); or a first-degree relative with OCD or tic disorder. Exclusion criteria for all included: any MRI contraindications; major medical or neurological disease/head trauma; pregnancy; acute suicidality; or an intelligence quotient (IQ) below 80 (24)^1^. Participants provided written informed consent and local ethical committees at each site approved the study.

### Measures

Trained raters (27) measured the severity of obsessive-compulsive symptoms using the Y-BOCS (33), the severity of aggressive, sexual/religious, symmetry and contamination symptoms using the Dimensional Yale-Brown Obsessive-Compulsive Scale (DY-BOCS) (34), the severity of anxiety symptoms using the Hamilton Anxiety Rating Scale (HAM-A) (35), and the severity of depressive symptoms using the Hamilton Depression Rating Scale (HAM-D) (36),

### MRI acquisition and preprocessing

As previously reported in detail elsewhere (25), a structural T1-weighted image was acquired using magnetization-prepared rapid acquisition gradient-echo imaging according to the ADNI-3 protocol. Eyes-closed rs-fMRI was performed for 10 minutes using gradient echo planar imaging and a field map with opposite phase encoding directions (See Table S1 for details).

Structural images were processed in FreeSurfer (version 7.1.1) (37) and fMRI data were preprocessed using fMRIPrep (version 20.2.3, see Supplemental Material for details) (38). The functional images were corrected for susceptibility-induced distortions, co-registered to the structural image followed by motion estimation and slice time correction, and the first three volumes of the functional images were discarded to ensure stable spin magnetization. Images were then registered to “grayordinate” space (MNINonLinear/fsLR32k) using Ciftify (39), which combines a cortical surface-based framework with the volumetric subcortical regions, providing greater anatomical specificity than traditional volumetric representations of the brain (40). Here, cortical gray matter time series were sampled onto the cortical surface using the MSMSulc algorithm based on each participant’s reconstructed cortex using FreeSurfer. Subcortical time series were registered to MNI space using FSL FNIRT.

Participants with mean framewise displacement (FD) >0.5mm were excluded from further analysis (41). Functional MRI data were denoised using nuisance regression consisting of mean, squares, temporal derivatives, and squares of derivatives of cerebrospinal fluid and white matter signal and motion components from ICA-AROMA, followed by linear detrending (41). Time courses were bandpass filtered to between 0.009 Hz and 0.08 Hz and minimal smoothing was performed through regionally constrained 2mm Gaussian kernels. ComBat was used to adjust for differences in rs-fMRI measures between sites while retaining variance related to age, sex, years of education, and diagnosis of OCD for every vertex/voxel for seed-based analyses, for connectivity values for between/within network FC and global network topology, and for connectivity matrices for level for NBS (42).

### Seed-based FC

CSTC circuits were operationalized using bilateral key regions of each circuit as seeds in accordance with previous studies (9, 20). The ventral cognitive circuit was probed using the bilateral ventral caudate, the dorsal cognitive circuit using the bilateral dorsal caudate, the sensorimotor circuit using the posterior putamen, the frontolimbic circuit using the amygdala, and the affective circuit using the bilateral nucleus accumbens. All seeds were based on the Brainnetome atlas which separates the anterior and posterior putamen and dorsal and ventral caudate based on multimodal connectivity (43). R-to-z Fisher-transformed correlation maps between the respective seed regions and the rest of the brain were calculated. The maps were then split up into the left surface, right surface and subcortex, submitted to univariate general linear models in Permutation Analysis of Linear Models (PALM) (44), followed by combining the thresholded FC maps of the left surface, right surface and subcortex for visualization.

### Within and between subnetwork FC

Following procedures by ENIGMA-OCD (13), seven subnetworks (visual, somatomotor, dorsal attention [DAN], ventral attention [VAN], limbic, frontoparietal [FPN], and default mode [DMN]) were defined using the Schaefer 400 cortical parcellations (45) and 36 subcortical parcellations defined using the Brainnetome atlas (43). Connectivity matrices were constructed from the extracted time courses using Pearson correlation coefficients, which were subsequently R-to-z Fisher-transformed. Mean FC was calculated for connections within each subnetwork (within-subnetwork FC) and connections between each pair of subnetworks (between-subnetwork FC).

### Network-based Statistic

NBS is a commonly used method for finding connections that are maximally different between groups or related to a variable of interest (6). NBS first performs mass univariate testing of all connections at a threshold of t≥3.1, followed by cluster-wise correction based on the number of suprathreshold connections (6).

### Global network topology

We calculated global efficiency and modularity from the full weighted network of 400 cortical and 36 subcortical regions using the Brain Connectivity Toolbox (7). Global efficiency measures the degree of functional integration or how easily information can cross throughout the network, while modularity is the degree to which the network can be divided into functionally distinct communities (7).

### Statistical analysis

The analysis plan was pre-registered at the Open Science Foundation (https://osf.io/b3vz5). Following that plan, we used separate models for comparing OCD vs HC and comparing subgroups and associations with clinical characteristic within OCD. This included early versus late onset of OCD, prior use of SSRI/SNRI, current comorbid anxiety disorders, current comorbid major depressive disorder (MDD), Y-BOCS, D-YBOCS, HAM-A, and HAM-D. All models included a single independent variable of interest, except for the four DY-BOCS subscales, which were entered in the same model. All models were further adjusted for sex, age, and years of education as covariates. Univariate general linear models for seed-based FC were performed in PALM (version 119) using Threshold-Free Cluster Enhancement (TFCE) with 5,000 permutations and significance set to family-wise error (FWE) p<0.05. Univariate general linear models of subnetwork-based FC and graph FC measures were performed in R (version 4.2.1) using the lmPerm package (version 2.1.0) and 10,000 permutations, with significance set to False Discovery Rate (FDR) p<0.05. Finally, network-based analyses in the NBS toolbox (version 1.2) used the conventional t≥3.1 as the initial threshold and FWEp<.05 as the cluster-wise threshold and 5000 permutations (6). We also explored a higher threshold of t≥4 to examine the breakdown of significant clusters.

### Data Availability and Reproducibility

Upon study completion, data from this study will be submitted to the NIMH Data Archive, an NIH-funded data repository.

## Results

### Participants characteristics

The initial sample consisted of 281 adults with OCD and 265 HC. Twelve participants (n=7 OCD; n=5 HC) were excluded after enrollment due to withdrawal of consent, not completing any procedures, or later meeting study exclusion criteria. This left 274 OCD and 260 HC participants, of whom 11 (n=7 OCD, n=4 HC) did not complete the MRI with usable data.. As a result, MRI data were available for 523 participants (n=267 OCD; 256 HC). We additionally excluded six participants due to mean FD exceeding 0.5mm (n=4 OCD) or because of enlarged ventricles that impeded accurate registration (n=2 HC). Thus, the final analyzed sample included 263 OCD and 254 HC.

The OCD and HC groups were well matched in terms of sex, age, and mean FD, while HC showed significantly more years of education and higher IQ (Table 1). We compared subgroups of OCD based on age of onset, prior use of SSRI/SNRI, and comorbid anxiety disorders or MDD. Early onset of OCD was associated with significantly younger age, higher IQ, more comorbid anxiety disorders, and fewer individuals being naïve to antipsychotics and CBT (Table S2). Prior use of SSRI/SNRI (versus being medication naïve) was associated with significantly older age, higher Y-BOCS, HAM-A and HAM-D scores, more comorbid MDD, and fewer individuals being naïve to benzodiazepines, antipsychotics, and CBT (Table S3). Having a comorbid anxiety disorder was significantly associated with early-onset OCD, more severe HAM-A, HAM-D, and more comorbid MDD (Table S4). Having comorbid MDD was significantly associated with being female, more severe Y-BOCS, HAM-A, HAM-D, harm & aggression symptoms, and previous use of SSRI/SNRI (Table S5).

**Table 1.**
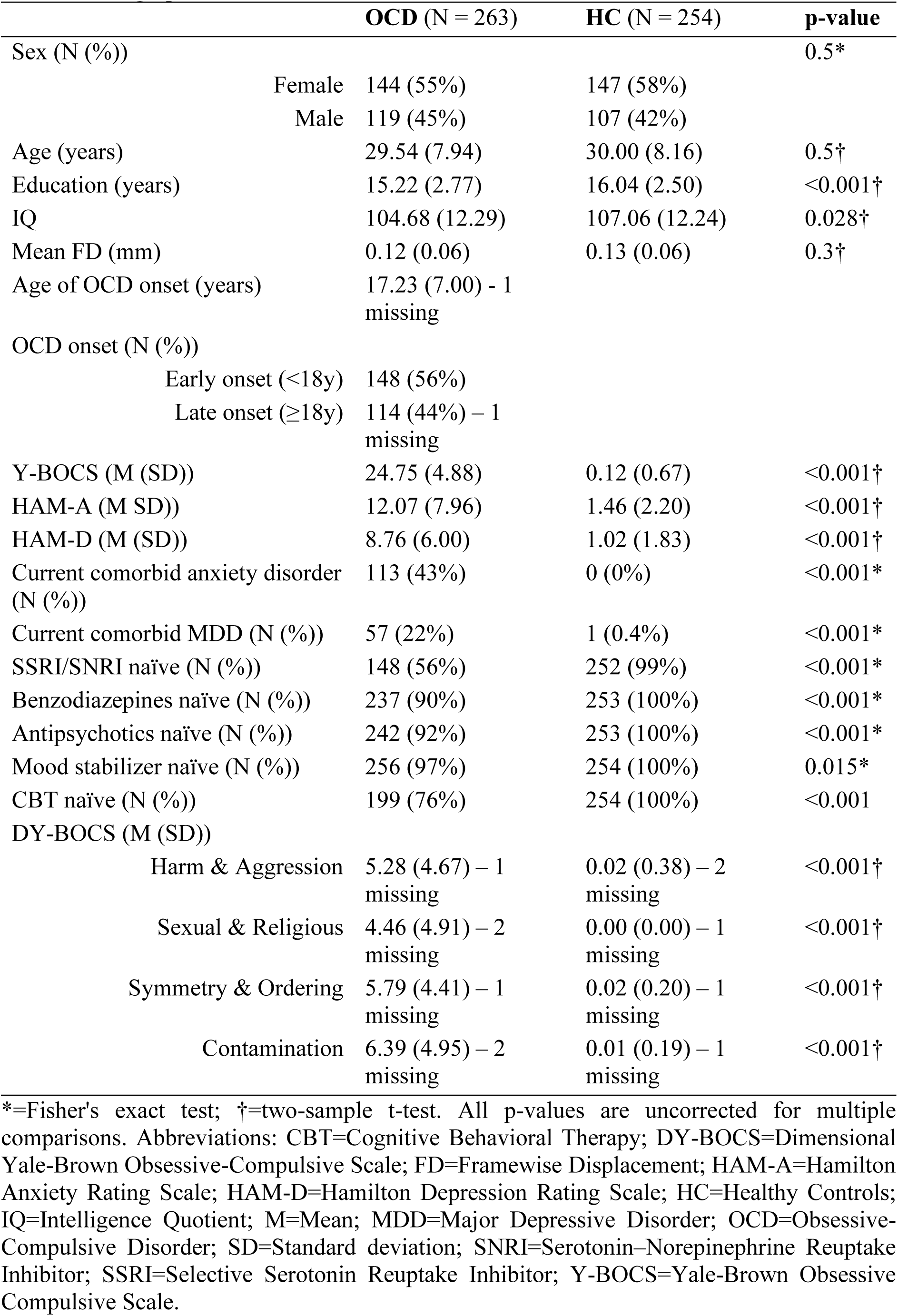
Demographic and clinical characteristics in OCD vs HC.

### Seed-based FC

We found no significant case-control differences in connectivity of the bilateral ventral caudate nucleus (ventral cognitive CSTC), dorsal caudate nucleus (dorsal cognitive CSTC), posterior putamen (sensorimotor CSTC), or amygdala (frontolimbic CSTC) circuits. We found significantly weaker FC between the nucleus accumbens (affective circuit) and the left cerebellum (Crus II region) in individuals with OCD versus HC (Figure 1, t=4.3, FWEp=.005, MNI XYZ=-12, –86, –38).

**Figure 1.**
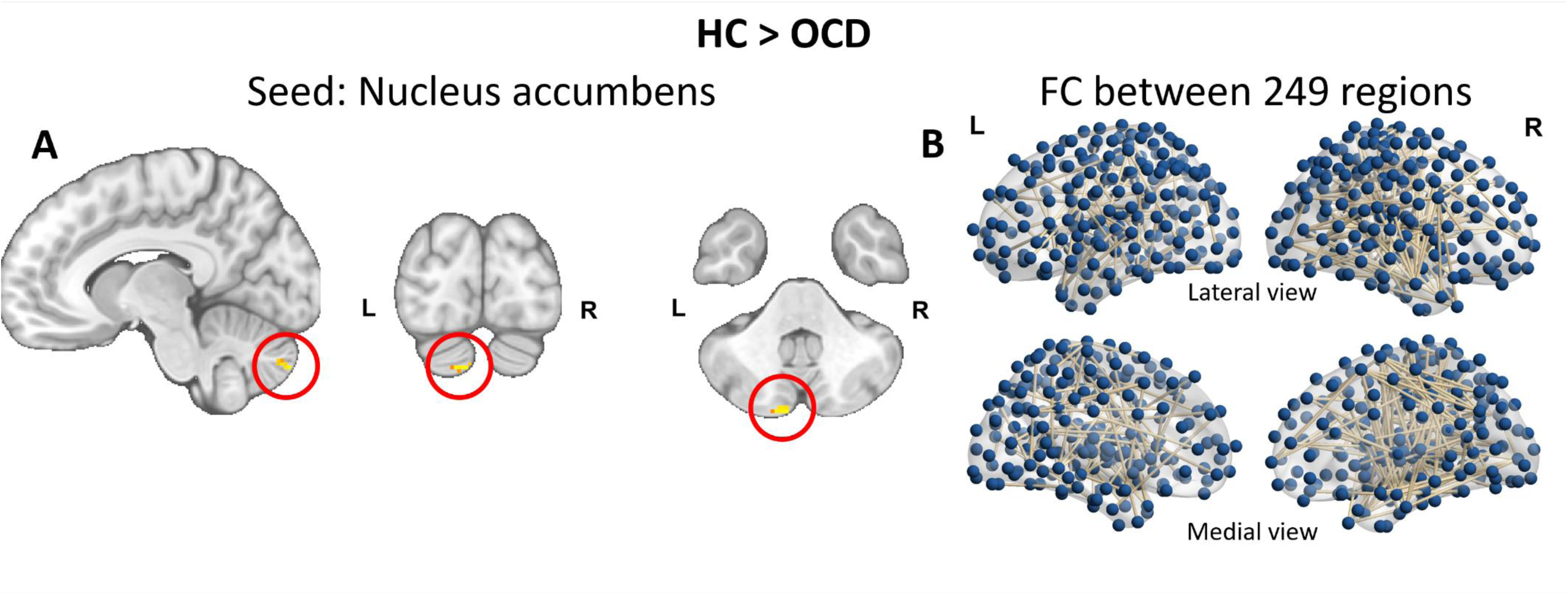
Differences in FC between OCD and HC. Figure legend: Panel A: Seed-based analyses showed that individuals with OCD had weaker FC compared to HC between the posterior putamen and left cerebellum Crus II in yellow voxels highlighted in the red circles. Panel B: Network-based statistic found that individuals with OCD showed widespread weaker FC compared to HC, where lines indicating significantly weaker connections between regions marked by blue dots.

Within the OCD group, we found significantly weaker FC between the posterior putamen (sensorimotor CSTC circuit) and the right thalamus in those with comorbid MDD compared to those without MDD (t=4.3, FWEp=.012, MNI XYZ=14, –26, 4), positive correlations between the severity of sexual & religious symptoms and FC between the amygdala (frontolimbic circuit) and right Crus II region of the cerebellum (t=4.7, FWEp=.009, MNI XYZ=16, –84, –36), as well as FC between the posterior putamen (sensorimotor circuit) and an extended cluster including left precentral, postcentral, lateral occipital, lingual, superior temporal, and supramarginal gyri (t=4.9, FWEp<.001)(Table 2 and Figure 2). We also found a positive correlation between HAM-A score and FC between the ventral caudate nucleus (ventral cognitive circuit) and the right precuneus (t=4.6, FWEp=.013), right angular gyrus (t=3.7, FWEp=.016), and right inferior parietal gyri (t=3.7, FWEp=.016) (Table 2 and Figure 2).

**Figure 2.**
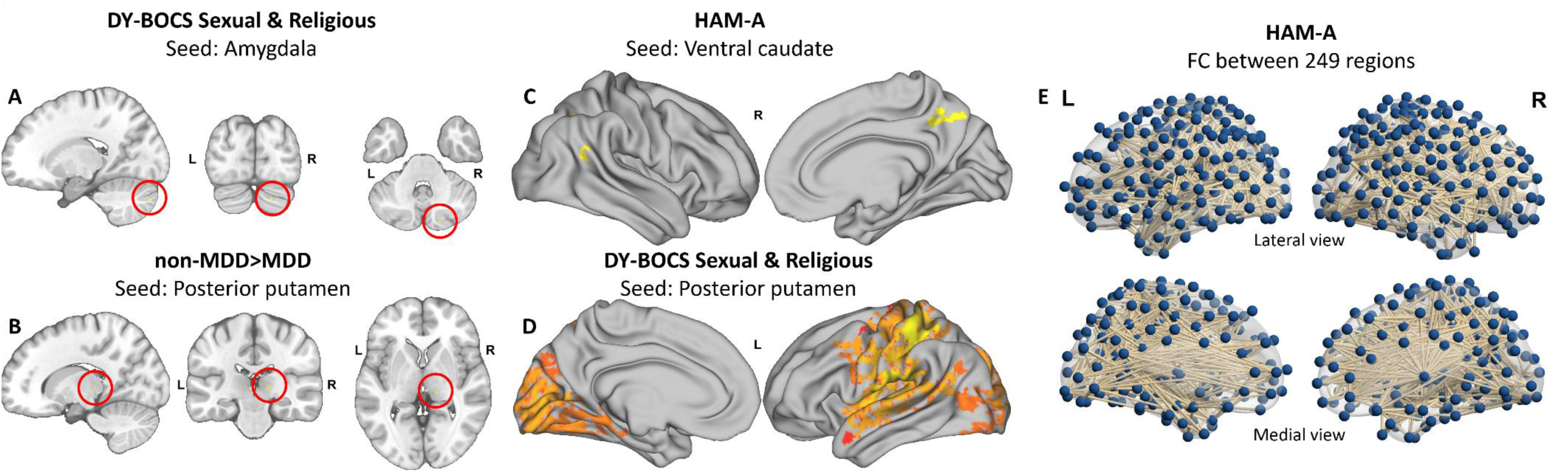
Associations between FC and clinical characteristics in OCD. Figure legend: Panel A: Seed-based analyses found a positive correlation between sexual & religious symptoms and FC between the amygdala and right cerebellum Crus II (highlighted in red circles). Panel B: Seed-based analyses found lower FC between the posterior putamen and right thalamus in non-depressed OCD (indicated by red-to-yellow colors). Panel C: Seed-based analyses found a positive correlation between HAM-A symptoms and FC between the ventral caudate and precuneus, angular gyrus, and inferior parietal gyrus (indicated by red-to-yellow colors). Panel D: Seed-based analyses found a positive correlation between sexual & religious symptoms and FC between the posterior putamen and extended sensorimotor areas. Panel E: Network-based statistic showed widespread positive correlation between HAM-A symptoms and FC (with lines indicating significantly weaker connections between regions marked by blue dots).

**Table 2.** Significant findings when comparing OCD (n=263) vs HC (n=254) and investigating clinical characteristics within OCD.

| Analysis | Circuit/subnetwork(s) | Regions | Contrast | Statistic | Corrected p |
| --- | --- | --- | --- | --- | --- |
| Seed-based | Affective | Nucleus accumbens - L<br>cerebellum Crus II | HC>OCD | t=4.3 | =.005 |
| Seed-based | Sensorimotor | Posterior putamen – R thalamus | non-MDD>MDD in OCD | t=4.3 | =.012 |
| Seed-based | Ventral cognitive | Ventral caudate nucleus – R<br>precuneus/R inferior parietal<br>cortex | Positive correlation with HAM-A in<br>OCD | t≥3.7 | ≤.016 |
| Seed-based | Frontolimbic | Amygdala - R cerebellum Crus II | Positive correlation with DY-BOCS<br>sexual & religious symptoms | t=4.7 | =.009 |
| Seed-based | Sensorimotor | Posterior putamen – extended<br>motor and visual cortex | Positive correlation with DY-BOCS<br>sexual & religious symptoms | t=4.9 | <.001 |
| Subnetwork-based | VAN-limbic | N/A | SSRI/SNRI naive<previous<br>SSRI/SNRI use | t=3.8 | <.001 |
| Subnetwork-based | VAN-DMN | N/A | SSRI/SNRI naive<previous<br>SSRI/SNRI use | t=3.3 | =.001 |
| NBS | All | 249 regions | HC>OCD | t≥3.1 | =.023 |
| NBS | All | 407 regions | Positive correlation with HAM-A in<br>OCD | t≥3.1 | =.035 |
| Global<br>efficiency | All | N/A | Negative correlation with DY-BOCS<br>Contamination in OCD | r=-0.1 | =.04 |

Abbreviations: DMN=Default Mode Network; DY-BOCS=Dimensional Yale-Brown Obsessive-Compulsive Scale; HAM-A=Hamilton Anxiety Rating Scale; HC=Healthy Controls; L=Left; MDD=Major Depressive Disorder; NBS=Network-based Statistic; OCD=Obsessive-Compulsive Disorder; R=Right; SNRI=Serotonin–Norepinephrine Reuptake Inhibitor; SSRI=Selective Serotonin Reuptake Inhibitor; VAN=Ventral Attention Network.

### Within and between subnetwork FC

We found no significant case-control difference in connectivity of the sensorimotor subnetwork. Within the OCD group we found weaker FC between the VAN and limbic subnetwork (t=3.82, FDRp<.001, d=0.47) and between the VAN and DMN (t=3.26, FDRp=0.01, d=0.41) in those with prior exposure to SSRI/SNRI versus those who were medication naive (Table 2).

### Network-based Statistic

We found a widespread pattern of lower FC in OCD compared to HC, involving 249 out of a total of 436 regions (Figure 1, FWEp<.05, t≥3.1). Further analyses at a stricter threshold of t≥4 revealed that this could be broken down into an occipital-parietal subnetwork of six regions, and a larger subnetwork spanning 15 parietal, temporal, subcortical, and prefrontal regions.

Within the OCD group, we found positive associations between HAM-A scores and FC of a widespread network spanning 409 regions (FWEp<.05, t≥3.1). A stricter threshold of t≥4 did not reveal further subnetworks but restricted the findings to 100 significant connections spanning all lobes of the brain.

### Global network topology

We found no significant differences between OCD and HC or associations within the OCD group, with the exception that more severe contamination symptoms were associated with lower global efficiency in OCD (r=-0.12, FDRp=.04, Table 2, Figure 2).

## Discussion

Using four complementary analytical approaches in the largest study to date with harmonized data collection across five continents and state-of-the-art processing, we investigated alterations in FC between currently unmedicated adults with OCD and HC, as well as associations between FC and clinical characteristics within OCD. This expands upon our recent reports of altered structural connectivity (28) and gray matter morphology (29) in the same sample. We found widespread hypoconnectivity in OCD versus HC using the NBS approach, as well as hypoconnectivity between the nucleus accumbens (considered a hub region in the affective CSTC) and the left Crus II of the cerebellum in the seed-based analyses. In the OCD group, we also observed significant associations of FC with prior use of SSRI/SNRI, comorbid depression, and the severity of anxiety and contamination symptoms. These findings underscore the importance of considering the heterogeneous nature of OCD and using multiple analytic approaches when investigating resting-state FC.

Our finding of widespread hypoconnectivity in OCD supports previous findings from meta-analyses of seed-based studies (4, 5), previous studies using NBS in OCD (46, 47), as well as the mega-analysis from the ENIGMA-OCD consortium (13). Our finding of hypoconnectivity between the nucleus accumbens and Crus II from the seed-based analysis may be related to increased sensitivity to possible threats and emotion regulation difficulties in OCD. This is supported by the role of the Crus II in tasks involving emotional self-experiences as well as explicit and implicit emotional processing (48). Furthermore, previous studies have found OCD-related hypoconnectivity of multiple cerebellar regions thought to be involved in emotional, cognitive, and sensorimotor processes (30, 46, 49, 50). In sum, our findings support that OCD is related to hypoconnectivity between multiple resting-state networks, but that the extent of this finding is influenced by the analytical approach. They support conceptualizing OCD and other mental illnesses as network disorders of the brain, rather than only being related to isolated brain regions or circuits (51). Our findings may also aid understanding the effects of interventions modulating the brain, such as repetitive transcranial magnetic stimulation (rTMS) (52). Notably, rTMS to the dorsolateral, dorsomedial, and pre-supplementary motor cortices have all found to be efficacious in OCD, possibly due to affecting widespread and disorder-related hypoconnectivity (53, 54).

Our sample size and detailed clinical phenotyping allowed for investigating how clinical characteristics in OCD relate to FC. This uncovered that even previous SSRI/SNRI use is associated with weaker VAN-limbic subnetwork and VAN-DMN FC in OCD. The VAN is central to detecting and filtering salient stimuli, processes which have repeatedly been implicated in OCD (1–3). This is supported by experiments showing that SSRI/SNRI administration results in reduced resting-state FC even in healthy volunteers with no effects on mood or subjective state (55, 56). This dovetails with previous functional and structural neuroimaging studies of current medication use in OCD and further suggests that prior SSRI/SNRI should be considered, even when participants are not currently taking medication (OCD) (1–3). However, separating the effects of SSRI/SNRI on FC versus other factors is not possible with a cross-sectional study design, and it is worth highlighting that individuals with OCD previously using SSRI/SNRIs in the present study were older, had more severe obsessive-compulsive, anxiety, and depressive symptoms, higher rates of comorbid MDD, and previous use of other pharmacological and psychological treatments.

We found that individuals with OCD with more severe anxiety symptoms showed stronger FC both widespread across the brain (based on the NBS analyses) and between the ventral caudate nucleus (ventral cognitive circuit), right precuneus, and the inferior parietal cortex (based on the seed-based approach). This mirrors previous findings in social and generalized anxiety disorders, possibly suggesting that anxiety is linked to more crosstalk between multiple circuits and subnetworks as a function of overactive threat monitoring and worry (57–59). At the same time, individuals with OCD and comorbid MDD showed weaker FC between the bilateral posterior putamen (sensorimotor circuit) and the right thalamus, which are key areas linked to sensory symptoms and habit formation in OCD (3).

When investigating the severity of specific obsessive-compulsive symptom dimensions, we found that more severe sexual and religious symptoms were linked to stronger FC between the posterior putamen and other regions within the sensorimotor circuit extending to areas of the VAN and DAN, and stronger FC between the frontolimbic circuit and right Crus II of the cerebellum. Notably, similar associations between sexual & religious symptoms and altered recruitment of the frontolimbic and sensorimotor regions have also emerged in previous symptom provocation studies, which may reflect both increased threat processing and readiness for a behavioral response (60). Lastly, we found that more severe contamination symptoms were linked to less global efficiency in OCD, which is a novel finding in the literature. This suggests that the role of different symptom dimensions should be assessed in future studies of functional global efficiency, as comparisons between individuals with OCD and HC have yielded inconsistent findings (15, 19, 61).

A comparison of the present findings on FC with those found using diffusion-weighted imaging (28) and gray matter morphology in the Global OCD study (29) reveals both overlap and modality-specific findings. Both rs-fMRI and gray matter morphology-derived measures showed correlations with the severity of anxiety symptoms, the severity of sexual and religious symptoms, and comorbid depression in OCD. Individuals with OCD and greater severity of anxiety and sexual and religious symptoms tended to more strongly resemble healthy controls, as characterized by widespread greater FC, larger thalamus volume and hippocampal volume. Meanwhile, individuals with OCD and comorbid depression showed both smaller surface area in several regions and weaker FC between the thalamus and sensorimotor circuit. In contrast, alterations in structural connectivity were most pronounced in late-onset cases, which was not the case for FC or gray matter morphology. Interestingly, SSRI/SNRI naïve participants showed less regional white matter fractional anisotropy compared to participants previously using SSRI/SNRI, contrasting the present findings (28). These divergent results suggest that OCD-related alterations in functional and structural connectivity are likely influenced by different clinical characteristics, including age of onset, medication history, comorbid disorders or the severity of comorbid depressive and anxiety symptoms, and the type of obsessive-compulsive symptoms.

Our study has several strengths including a large, well-characterized sample that included unmedicated OCD participants and matched healthy controls, recruitment across five global sites increasing generalizability of the findings, and use of multiple approaches to analyze resting-state FC, which uncovered both distinct and common findings. It also has several limitations. First, using multiple approaches leads to an increased number of tests and a higher likelihood of false positive findings. However, we consider this a reasonable tradeoff given that the different analytic approaches are sensitive to different aspects of functional brain organization and that there is currently no single optimal method for investigating resting-state FC covering all aspects of mental disorders. We suggest that future studies also pre-register hypotheses and methodological choices and use multiple approaches where applicable. Second, there was a significant difference between years of education and IQ in OCD compared to HC (potentially reflecting the impact of OCD on educational attainment and cognitive function), as well as differences in demographic and clinical variables between subgroups with OCD. However, the size of these differences was generally small to moderate. (39, 40).

In conclusion, the present study indicates that individuals with OCD have both widespread and region-specific hypoconnectivity compared to HC. We also show that previous SSRI/SNRI use, comorbid MDD, severity of anxiety symptoms, and contamination, sexual, and religious symptoms are linked to interindividual variation in OCD. Finally, we demonstrate that using multiple levels of analysis uncovers both shared and distinct results, highlighting the need to conduct and report analyses from multiple perspectives in future studies.

## Acknowledgements

The authors would like to thank all participants of this study and the following study team members from each site for their vital contributions to this project: Marines Joaquim, RN, Maria Alice de Mathis, PhD and Daniel L.C. Costa, MD PhD (Brazil); Anton J.L.M. van Balkom, MD PhD, Neeltje Batelaan, MD PhD (Netherlands); Loche Manuel, Hons, Clara Marincowitz, MSc (South Africa); Rachel Middleton B.A., Gabrielle R. Messner B.A., Gabriella Restifo-Bernstein, B.A., Sarah Rose, B.A., Yael R. Stovetsky B.A. (U.S.A). We also thank Page Van Meter, Ph.D., for overseeing all data management for this study. Preliminary results were presented in the form of a poster at the Society of Biological Psychiatry annual meeting in Toronto; Canada, April 24-26, 2025. A preprint of this article was deposited at medRxiv (URL) prior to submission to the journal.

## Footnotes

1 Minor exceptions were made in 13 cases (6 OCD, 7HC) with IRB notice.

## References

1. Stein DJ, Costa DLC, Lochner C, Miguel EC, Reddy YCJ, Shavitt RG, et al. Obsessive–compulsive disorder. Nat Rev Dis Primers. 2019;5(1):1–21.

2. Thorsen AL, van den Heuvel OA. Neuroanatomy of Obsessive-Compulsive and Related Disorders. In: Tolin D, editor. The Oxford Handbook of Obsessive-Compulsive and Related Disorders: Oxford University Press; 2023. p. 143–173.

3. Shephard E, Stern ER, van den Heuvel OA, Costa DLC, Batistuzzo MC, Godoy PBG, et al. Toward a neurocircuit-based taxonomy to guide treatment of obsessive-compulsive disorder. Mol Psychiatry. 2021;26(9):4583–4604.

4. Gursel DA, Avram M, Sorg C, Brandl F, Koch K. Frontoparietal areas link impairments of large-scale intrinsic brain networks with aberrant fronto-striatal interactions in OCD: a meta-analysis of resting-state functional connectivity. Neurosci Biobehav Rev. 2018;87:151–160.

5. Liu J, Cao L, Li H, Gao Y, Bu X, Liang K, et al. Abnormal resting-state functional connectivity in patients with obsessive-compulsive disorder: A systematic review and meta-analysis. Neurosci Biobehav Rev. 2022;135:104574.

6. Zalesky A, Fornito A, Bullmore ET. Network-based statistic: identifying differences in brain networks. Neuroimage. 2010;53(4):1197–207.

7. Rubinov M, Sporns O. Complex network measures of brain connectivity: uses and interpretations. Neuroimage. 2010;52(3):1059–69.

8. de Vries FE, de Wit SJ, van den Heuvel OA, Veltman DJ, Cath DC, van Balkom A, et al. Cognitive control networks in OCD: A resting-state connectivity study in unmedicated patients with obsessive-compulsive disorder and their unaffected relatives. World J Biol Psychiatry. 2017:1–13.

9. Harrison BJ, Soriano-Mas C, Pujol J, Ortiz H, Lopez-Sola M, Hernandez-Ribas R, et al. Altered corticostriatal functional connectivity in obsessive-compulsive disorder. Arch Gen Psychiatry. 2009;66(11):1189–200.

10. Moreira PS, Marques P, Soriano-Mas C, Magalhaes R, Sousa N, Soares JM, et al. The neural correlates of obsessive-compulsive disorder: a multimodal perspective. Transl Psychiatry. 2017;7(8):e1224.

11. Vaghi MM, Vértes PE, Kitzbichler MG, Apergis-Schoute AM, van der Flier FE, Fineberg NA, et al. Specific Frontostriatal Circuits for Impaired Cognitive Flexibility and Goal-Directed Planning in Obsessive-Compulsive Disorder: Evidence From Resting-State Functional Connectivity. Biol Psychiatry. 2017;81(8):708–717.

12. Posner J, Song I, Lee S, Rodriguez CI, Moore H, Marsh R, et al. Increased functional connectivity between the default mode and salience networks in unmedicated adults with obsessive-compulsive disorder. Hum Brain Mapp. 2017;38(2):678–687.

13. Bruin WB, Abe Y, Alonso P, Brem S, Walitza S, ENIGMA-OCD Working Group, et al. The functional connectome in obsessive-compulsive disorder: resting-state mega-analysis and machine learning classification for the ENIGMA-OCD consortium. Mol Psychiatry. 2023.

14. Jung WH, Yucel M, Yun JY, Yoon YB, Cho KI, Parkes L, et al. Altered functional network architecture in orbitofronto-striato-thalamic circuit of unmedicated patients with obsessive-compulsive disorder. Hum Brain Mapp. 2017;38(1):109–119.

15. Shin DJ, Jung WH, He Y, Wang J, Shim G, Byun MS, et al. The effects of pharmacological treatment on functional brain connectome in obsessive-compulsive disorder. Biol Psychiatry. 2014;75(8):606–14.

16. Anticevic A, Hu S, Zhang S, Savic A, Billingslea E, Wasylink S, et al. Global resting-state functional magnetic resonance imaging analysis identifies frontal cortex, striatal, and cerebellar dysconnectivity in obsessive-compulsive disorder. Biol Psychiatry. 2014;75(8):595–605.

17. Li X, Li H, Jiang X, Li J, Cao L, Liu J, et al. Characterizing multiscale modular structures in medication-free obsessive-compulsive disorder patients with no comorbidity. Hum Brain Mapp. 2022;43(7):2391–2399.

18. Beucke JC, Sepulcre J, Talukdar T, Linnman C, Zschenderlein K, Endrass T, et al. Abnormally high degree connectivity of the orbitofrontal cortex in obsessive-compulsive disorder. JAMA Psychiatry. 2013;70(6):619–29.

19. Thorsen AL, Vriend C, de Wit SJ, Ousdal OT, Hagen K, Hansen B, et al. Effects of Bergen 4-day treatment on resting-state graph features in obsessive-compulsive disorder. Biological Psychiatry: Cognitive Neuroscience and Neuroimaging. 2021;6(10):973–982.

20. Harrison BJ, Pujol J, Cardoner N, Deus J, Alonso P, Lopez-Sola M, et al. Brain corticostriatal systems and the major clinical symptom dimensions of obsessive-compulsive disorder. Biol Psychiatry. 2013;73(4):321–8.

21. Fan J, Zhong M, Zhu X, Gan J, Liu W, Niu C, et al. Resting-state functional connectivity between right anterior insula and right orbital frontal cortex correlate with insight level in obsessive-compulsive disorder. Neuroimage Clin. 2017;15:1–7.

22. Cao L, Li H, Hu X, Liu J, Gao Y, Liang K, et al. Distinct alterations of amygdala subregional functional connectivity in early– and late-onset obsessive-compulsive disorder. J Affect Disord. 2022;298(Pt A):421–430.

23. Tadayonnejad R, Deshpande R, Ajilore O, Moody T, Morfini F, Ly R, et al. Pregenual anterior cingulate dysfunction associated with depression in OCD: an integrated multimodal fMRI/(1)h MRS study. Neuropsychopharmacology. 2018;43(5):1146–1155.

24. Simpson HB, van den Heuvel OA, Miguel EC, Reddy YCJ, Stein DJ, Lewis-Fernández R, et al. Toward identifying reproducible brain signatures of obsessive-compulsive profiles: rationale and methods for a new global initiative. BMC Psychiatry. 2020;20(1):68.

25. Pouwels PJW, Vriend C, Liu F, de Joode NT, Otaduy MCG, Pastorello B, et al. Global multi-center and multi-modal magnetic resonance imaging study of obsessive-compulsive disorder: Harmonization and monitoring of protocols in healthy volunteers and phantoms. Int J Methods Psychiatr Res. 2022:e1931.

26. Batistuzzo MC, Sheshachala K, Alschuler DM, Hezel DM, Lewis-Fernández R, de Joode NT, et al. Cross-national harmonization of neurocognitive assessment across five sites in a global study. Neuropsychology. 2022.

27. Shavitt RG, Sheshachala K, Hezel DM, Wall MM, Balachander S, Lochner C, et al. Measurement fidelity of clinical assessment methods in a global study on identifying reproducible brain signatures of obsessive-compulsive disorder. Neuropsychology. 2023;37(3):330–343.

28. Vriend C, de Joode NT, Pouwels PJW, Liu F, Otaduy MCG, Pastorello B, et al. Age of onset of obsessive-compulsive disorder differentially affects white matter microstructure. Mol Psychiatry. 2024;29(4):1033–1045.

29. de Joode NT, Vriend C, Pouwels PJW, Liu F, Otaduy MCG, Pastorello B, et al. Cortical and subcortical structural alterations in obsessive-compulsive disorder: relationships between morphology and clinical profiles in the Global OCD study. medRxiv. 2025:2025.07.23.25332068.

30. Xu T, Zhao Q, Wang P, Fan Q, Chen J, Zhang H, et al. Altered resting-state cerebellar-cerebral functional connectivity in obsessive-compulsive disorder. Psychol Med. 2019;49(7):1156–1165.

31. Yeo BT, Krienen FM, Sepulcre J, Sabuncu MR, Lashkari D, Hollinshead M, et al. The organization of the human cerebral cortex estimated by intrinsic functional connectivity. J Neurophysiol. 2011;106(3):1125–65.

32. First MB, Williams JB, Karg RS, Spitzer RL. Structured Clinical Interview for DSM-5 Disorders. Arlington, VA: American Psychiatric Association; 2016 p.

33. Goodman WK, Price LH, Rasmussen SA, Mazure C, Fleischmann RL, Hill CL, et al. The Yale-Brown Obsessive Compulsive Scale. I. Development, use, and reliability. Arch Gen Psychiatry. 1989;46(11):1006–11.

34. Rosario-Campos MC, Miguel EC, Quatrano S, Chacon P, Ferrao Y, Findley D, et al. The Dimensional Yale-Brown Obsessive-Compulsive Scale (DY-BOCS): an instrument for assessing obsessive-compulsive symptom dimensions. Mol Psychiatry. 2006;11(5):495–504.

35. Hamilton M. The assessment of anxiety states by rating. Br J Med Psychol. 1959;32(1):50–5.

36. Hamilton M. A rating scale for depression. J Neurol Neurosurg Psychiatry. 1960;23(1):56–62.

37. Fischl B. FreeSurfer. Neuroimage. 2012;62(2):774–81.

38. Esteban O, Markiewicz CJ, Blair RW, Moodie CA, Isik AI, Erramuzpe A, et al. fMRIPrep: a robust preprocessing pipeline for functional MRI. Nat Methods. 2019;16(1):111–116.

39. Dickie EW, Anticevic A, Smith DE, Coalson TS, Manogaran M, Calarco N, et al. Ciftify: A framework for surface-based analysis of legacy MR acquisitions. Neuroimage. 2019;197:818–826.

40. Coalson TS, Van Essen DC, Glasser MF. The impact of traditional neuroimaging methods on the spatial localization of cortical areas. Proc Natl Acad Sci U S A. 2018;115(27):E6356–e6365.

41. Parkes L, Fulcher B, Yücel M, Fornito A. An evaluation of the efficacy, reliability, and sensitivity of motion correction strategies for resting-state functional MRI. Neuroimage. 2018;171:415–436.

42. Radua J, Vieta E, Shinohara R, Kochunov P, Quidé Y, Green MJ, et al. Increased power by harmonizing structural MRI site differences with the ComBat batch adjustment method in ENIGMA. Neuroimage. 2020;218:116956.

43. Fan L, Li H, Zhuo J, Zhang Y, Wang J, Chen L, et al. The human brainnetome atlas: a new brain atlas based on connectional architecture. Cerebral cortex. 2016;26(8):3508–3526.

44. Winkler AM, Ridgway GR, Webster MA, Smith SM, Nichols TE. Permutation inference for the general linear model. Neuroimage. 2014;92:381–97.

45. Schaefer A, Kong R, Gordon EM, Laumann TO, Zuo XN, Holmes AJ, et al. Local-Global Parcellation of the Human Cerebral Cortex from Intrinsic Functional Connectivity MRI. Cereb Cortex. 2018;28(9):3095–3114.

46. Sha Z, Edmiston EK, Versace A, Fournier JC, Graur S, Greenberg T, et al. Functional Disruption of Cerebello-thalamo-cortical Networks in Obsessive-Compulsive Disorder. Biol Psychiatry Cogn Neurosci Neuroimaging. 2020;5(4):438–447.

47. Shi TC, Pagliaccio D, Cyr M, Simpson HB, Marsh R. Network-based functional connectivity predicts response to exposure therapy in unmedicated adults with obsessive-compulsive disorder. Neuropsychopharmacology. 2021;46(5):1035–1044.

48. Pierce JE, Thomasson M, Voruz P, Selosse G, Péron J. Explicit and Implicit Emotion Processing in the Cerebellum: A Meta-analysis and Systematic Review. Cerebellum. 2023;22(5):852–864.

49. Xing X, Jin L, Li Q, Yang Q, Han H, Xu C, et al. Modeling essential connections in obsessive-compulsive disorder patients using functional MRI. Brain Behav. 2020;10(2):e01499.

50. Zhang H, Wang B, Li K, Wang X, Li X, Zhu J, et al. Altered Functional Connectivity Between the Cerebellum and the Cortico-Striato-Thalamo-Cortical Circuit in Obsessive-Compulsive Disorder. Front Psychiatry. 2019;10:522.

51. Segal A, Tiego J, Parkes L, Holmes AJ, Marquand AF, Fornito A. Embracing variability in the search for biological mechanisms of psychiatric illness. Trends Cogn Sci. 2025;29(1):85–99.

52. Fitzsimmons S, Oostra E, Postma TS, van der Werf YD, van den Heuvel OA. Repetitive Transcranial Magnetic Stimulation-Induced Neuroplasticity and the Treatment of Psychiatric Disorders: State of the Evidence and Future Opportunities. Biol Psychiatry. 2024;95(6):592–600.

53. Fitzsimmons S, van der Werf YD, van Campen AD, Arns M, Sack AT, Hoogendoorn AW, et al. Repetitive transcranial magnetic stimulation for obsessive-compulsive disorder: A systematic review and pairwise/network meta-analysis. J Affect Disord. 2022;302:302–312.

54. Fitzsimmons S, Postma TS, van Campen AD, Vriend C, Batelaan NM, van Oppen P, et al. Transcranial Magnetic Stimulation-Induced Plasticity Improving Cognitive Control in Obsessive-Compulsive Disorder, Part I: Clinical and Neuroimaging Outcomes From a Randomized Trial. Biol Psychiatry. 2025;97(7):678–687.

55. McCabe C, Mishor Z. Antidepressant medications reduce subcortical-cortical resting-state functional connectivity in healthy volunteers. Neuroimage. 2011;57(4):1317–23.

56. McCabe C, Mishor Z, Filippini N, Cowen PJ, Taylor MJ, Harmer CJ. SSRI administration reduces resting state functional connectivity in dorso-medial prefrontal cortex. Mol Psychiatry. 2011;16(6):592–4.

57. Zhang X, Yang X, Wu B, Pan N, He M, Wang S, et al. Large-scale brain functional network abnormalities in social anxiety disorder. Psychol Med. 2023;53(13):6194–6204.

58. Yang X, Liu J, Meng Y, Xia M, Cui Z, Wu X, et al. Network analysis reveals disrupted functional brain circuitry in drug-naive social anxiety disorder. Neuroimage. 2019;190:213–223.

59. Porta-Casteràs D, Fullana MA, Tinoco D, Martínez-Zalacaín I, Pujol J, Palao DJ, et al. Prefrontal-amygdala connectivity in trait anxiety and generalized anxiety disorder: Testing the boundaries between healthy and pathological worries. J Affect Disord. 2020;267:211– 219.

60. Via E, Cardoner N, Pujol J, Alonso P, Lopez-Sola M, Real E, et al. Amygdala activation and symptom dimensions in obsessive-compulsive disorder. Br J Psychiatry. 2014;204(1):61–8.

61. Li X, Li H, Cao L, Liu J, Xing H, Huang X, et al. Application of graph theory across multiple frequency bands in drug-naïve obsessive-compulsive disorder with no comorbidity. J Psychiatr Res. 2022;150:272–278.

