## Supplemental Materials for "Resting-state functional connectivity alterations in obsessive-compulsive disorder: relationships between connectivity and clinical profiles in the Global OCD study"

### Boilerplate from fMRIPrep

Results included in this manuscript come from preprocessing performed using *fMRIPrep* 20.2.3 (Esteban, Markiewicz, et al. (2018); Esteban, Blair, et al. (2018); RRID:SCR_016216), which is based on *Nipype* 1.6.1 (Gorgolewski et al. (2011); Gorgolewski et al. (2018); RRID:SCR_002502).

*Anatomical data preprocessing*

A total of 1 T1-weighted (T1w) images were found within the input BIDS dataset.The T1-weighted (T1w) image was corrected for intensity non-uniformity (INU) with N4BiasFieldCorrection (Tustison et al. 2010), distributed with ANTs 2.3.3 (Avants et al. 2008, RRID:SCR_004757), and used as T1w-reference throughout the workflow. The T1w-reference was then skull-stripped with a *Nipype* implementation of the antsBrainExtraction.sh workflow (from ANTs), using OASIS30ANTs as target template. Brain tissue segmentation of cerebrospinal fluid (CSF), white-matter (WM) and gray-matter (GM) was performed on the brain-extracted T1w using fast (FSL 5.0.9, RRID:SCR_002823, Zhang, Brady, and Smith 2001). Brain surfaces were reconstructed using recon-all (FreeSurfer 6.0.1, RRID:SCR_001847, Dale, Fischl, and Sereno 1999), and the brain mask estimated previously was refined with a custom variation of the method to reconcile ANTs-derived and FreeSurfer-derived segmentations of the cortical gray-matter of Mindboggle (RRID:SCR_002438, Klein et al. 2017). Volume-based spatial normalization to two standard spaces (MNI152NLin6Asym, MNI152NLin2009cAsym) was performed through nonlinear registration with antsRegistration (ANTs 2.3.3), using brain-extracted versions of both T1w reference and the T1w template. The following templates were selected for spatial normalization: *FSL's MNI ICBM 152 non-linear 6th Generation Asymmetric Average Brain Stereotaxic Registration Model* [Evans et al. (2012), RRID:SCR_002823; TemplateFlow ID: MNI152NLin6Asym], *ICBM 152 Nonlinear Asymmetrical template version 2009c* [Fonov et al. (2009), RRID:SCR_008796; TemplateFlow ID: MNI152NLin2009cAsym],

*Functional data preprocessing*

For each of the 1 BOLD runs found per subject (across all tasks and sessions), the following preprocessing was performed. First, a reference volume and its skull-stripped version were generated using a custom methodology of *fMRIPrep*. A B0-nonuniformity map (or *fieldmap*) was estimated based on two (or more) echo-planar imaging (EPI) references with opposing phase-encoding directions, with 3dQwarp Cox and Hyde (1997) (AFNI 20160207). Based on the estimated susceptibility distortion, a corrected EPI (echo-planar imaging) reference was calculated for a more accurate co-registration with the anatomical reference. The BOLD reference was then co-registered to the T1w reference using bbregister (FreeSurfer) which implements boundary-based registration (Greve and Fischl 2009). Co-registration was configured with six degrees of freedom. Head-motion parameters with respect to the BOLD reference (transformation matrices, and six corresponding rotation and translation parameters) are estimated before any spatiotemporal filtering using mcflirt (FSL 5.0.9, Jenkinson et al. 2002). BOLD runs were slice-time corrected using 3dTshift from AFNI 20160207 (Cox and Hyde 1997, RRID:SCR_005927). The BOLD time-series were resampled onto the following surfaces (FreeSurfer reconstruction nomenclature): *fsnative, fsaverage5*. The BOLD time-series (including slice-timing correction when applied) were resampled onto their original, native space by applying a single, composite transform to correct for head-motion and susceptibility distortions. These resampled BOLD time-series will be referred to as *preprocessed BOLD in original space*, or just *preprocessed BOLD*. The BOLD time-series were resampled into standard space, generating a *preprocessed BOLD run in MNI152NLin6Asym space*. First, a reference volume and its skull-stripped version were generated using a custom methodology of *fMRIPrep*. Automatic removal of motion artifacts using independent component analysis (ICA-AROMA, Pruim et al. 2015) was performed on the *preprocessed BOLD on MNI space* time-series after removal of non-steady state volumes and spatial smoothing with an isotropic, Gaussian kernel of 6mm FWHM (full-width half-maximum). Corresponding "non-aggresively" denoised runs were produced after such smoothing. Additionally, the "aggressive" noise-regressors were collected and placed in the corresponding confounds file. Several confounding time-series were calculated based on the *preprocessed BOLD*: framewise displacement (FD), DVARS and three region-wise global signals. FD was computed using two formulations following Power (absolute sum of relative motions, Power et al. (2014)) and Jenkinson (relative root mean square displacement between affines, Jenkinson et al. (2002)). FD and DVARS are calculated for each functional run, both using their implementations in *Nipype* (following the definitions by Power et al. 2014). The three global signals are extracted within the CSF, the WM, and the whole-brain masks. Additionally, a set of physiological regressors were extracted to allow for component-based noise correction (*CompCor*, Behzadi et al. 2007). Principal components are estimated after high-pass filtering the *preprocessed BOLD* time-series (using a discrete cosine filter with 128s cut-off) for the two *CompCor* variants: temporal (tCompCor) and anatomical (aCompCor). tCompCor components are then calculated from the top 2% variable voxels within the brain mask. For aCompCor, three probabilistic masks (CSF, WM and combined CSF+WM) are generated in anatomical space. The implementation differs from that of Behzadi et al. in that instead of eroding the masks by 2 pixels on BOLD space, the aCompCor masks are subtracted a mask of pixels that likely contain a volume fraction of GM. This mask is obtained by dilating a GM mask extracted from the FreeSurfer's *aseg* segmentation, and it ensures components are not extracted from voxels containing a minimal fraction of GM. Finally, these masks are resampled into BOLD space and binarized by thresholding at 0.99 (as in the original implementation). Components are also calculated separately within the WM and CSF masks. For each CompCor decomposition, the *k* components with the largest singular values are retained, such that the retained components' time series are sufficient to explain 50 percent of variance across the nuisance mask (CSF, WM, combined, or temporal). The remaining components are dropped from consideration. The head-motion estimates calculated in the correction step were also placed within the corresponding confounds file. The confound time series derived from head motion estimates and global signals were expanded with the inclusion of temporal derivatives and quadratic terms for each (Satterthwaite et al. 2013). Frames that exceeded a threshold of 0.5 mm FD or 1.5 standardised DVARS were annotated as motion outliers. All resamplings can be performed with *a single interpolation step* by composing all the pertinent transformations (i.e. head-motion transform matrices, susceptibility distortion correction when available, and co-registrations to anatomical and output spaces). Gridded (volumetric) resamplings were performed using antsApplyTransforms (ANTs), configured with Lanczos interpolation to minimize the smoothing effects of other kernels (Lanczos 1964). Non-gridded (surface) resamplings were performed using mri_vol2surf (FreeSurfer).

Many internal operations of *fMRIPrep* use *Nilearn* 0.6.2 (Abraham et al. 2014, RRID:SCR_001362), mostly within the functional processing workflow. For more details of the pipeline, see the section corresponding to workflows in *fMRIPrep*'s documentation.

*Copyright Waiver*

The above boilerplate text was automatically generated by fMRIPrep with the express intention that users should copy and paste this text into their manuscripts *unchanged*. It is released under the CC0 license.

*References*

Abraham, Alexandre, Fabian Pedregosa, Michael Eickenberg, Philippe Gervais, Andreas Mueller, Jean Kossaifi, Alexandre Gramfort, Bertrand Thirion, and Gael Varoquaux. 2014. "Machine Learning for Neuroimaging with Scikit-Learn." *Frontiers in Neuroinformatics* 8. https://doi.org/10.3389/fninf.2014.00014.

Avants, B.B., C.L. Epstein, M. Grossman, and J.C. Gee. 2008. "Symmetric Diffeomorphic Image Registration with Cross-Correlation: Evaluating Automated Labeling of Elderly and Neurodegenerative Brain." *Medical Image Analysis* 12 (1): 26–41. https://doi.org/10.1016/j.media.2007.06.004.

Behzadi, Yashar, Khaled Restom, Joy Liau, and Thomas T. Liu. 2007. "A Component Based Noise Correction Method (CompCor) for BOLD and Perfusion Based fMRI." *NeuroImage* 37 (1): 90–101. https://doi.org/10.1016/j.neuroimage.2007.04.042.

Cox, Robert W., and James S. Hyde. 1997. "Software Tools for Analysis and Visualization of fMRI Data." *NMR in Biomedicine* 10 (4-5): 171–78. https://doi.org/10.1002/(SICI)1099-1492(199706/08)10:4/5<171::AID-NBM453>3.0.CO;2-L.

Dale, Anders M., Bruce Fischl, and Martin I. Sereno. 1999. "Cortical Surface-Based Analysis: I. Segmentation and Surface Reconstruction." *NeuroImage* 9 (2): 179–94. https://doi.org/10.1006/nimg.1998.0395.

Esteban, Oscar, Ross Blair, Christopher J. Markiewicz, Shoshana L. Berleant, Craig Moodie, Feilong Ma, Ayse Ilkay Isik, et al. 2018. "FMRIPrep." *Software*. Zenodo. https://doi.org/10.5281/zenodo.852659.

Esteban, Oscar, Christopher Markiewicz, Ross W Blair, Craig Moodie, Ayse Ilkay Isik, Asier Erramuzpe Aliaga, James Kent, et al. 2018. "fMRIPrep: A Robust Preprocessing Pipeline for Functional MRI." *Nature Methods*. https://doi.org/10.1038/s41592-018-0235-4.

Evans, AC, AL Janke, DL Collins, and S Baillet. 2012. "Brain Templates and Atlases." *NeuroImage* 62 (2): 911–22. https://doi.org/10.1016/j.neuroimage.2012.01.024.

Fonov, VS, AC Evans, RC McKinstry, CR Almli, and DL Collins. 2009. "Unbiased Nonlinear Average Age-Appropriate Brain Templates from Birth to Adulthood." *NeuroImage* 47, Supplement 1: S102. https://doi.org/10.1016/S1053-8119(09)70884-5.

Gorgolewski, K., C. D. Burns, C. Madison, D. Clark, Y. O. Halchenko, M. L. Waskom, and S. Ghosh. 2011. "Nipype: A Flexible, Lightweight and Extensible Neuroimaging Data Processing Framework in Python." *Frontiers in Neuroinformatics* 5: 13. https://doi.org/10.3389/fninf.2011.00013.

Gorgolewski, Krzysztof J., Oscar Esteban, Christopher J. Markiewicz, Erik Ziegler, David Gage Ellis, Michael Philipp Notter, Dorota Jarecka, et al. 2018. "Nipype." *Software*. Zenodo. https://doi.org/10.5281/zenodo.596855.

Greve, Douglas N, and Bruce Fischl. 2009. "Accurate and Robust Brain Image Alignment Using Boundary-Based Registration." *NeuroImage* 48 (1): 63–72. https://doi.org/10.1016/j.neuroimage.2009.06.060.

Jenkinson, Mark, Peter Bannister, Michael Brady, and Stephen Smith. 2002. "Improved Optimization for the Robust and Accurate Linear Registration and Motion Correction of Brain Images." *NeuroImage* 17 (2): 825–41. https://doi.org/10.1006/nimg.2002.1132.

Klein, Arno, Satrajit S. Ghosh, Forrest S. Bao, Joachim Giard, Yrjö Häme, Eliezer Stavsky, Noah Lee, et al. 2017. "Mindboggling Morphometry of Human Brains." *PLOS Computational Biology* 13 (2): e1005350. https://doi.org/10.1371/journal.pcbi.1005350.

Lanczos, C. 1964. "Evaluation of Noisy Data." *Journal of the Society for Industrial and Applied Mathematics Series B Numerical Analysis* 1 (1): 76–85. https://doi.org/10.1137/0701007.

Power, Jonathan D., Anish Mitra, Timothy O. Laumann, Abraham Z. Snyder, Bradley L. Schlaggar, and Steven E. Petersen. 2014. "Methods to Detect, Characterize, and Remove Motion Artifact in Resting State fMRI." *NeuroImage* 84 (Supplement C): 320–41. https://doi.org/10.1016/j.neuroimage.2013.08.048.

Pruim, Raimon H. R., Maarten Mennes, Daan van Rooij, Alberto Llera, Jan K. Buitelaar, and Christian F. Beckmann. 2015. "ICA-AROMA: A Robust ICA-Based Strategy for Removing Motion Artifacts from fMRI Data." *NeuroImage* 112 (Supplement C): 267–77. https://doi.org/10.1016/j.neuroimage.2015.02.064.

Satterthwaite, Theodore D., Mark A. Elliott, Raphael T. Gerraty, Kosha Ruparel, James Loughead, Monica E. Calkins, Simon B. Eickhoff, et al. 2013. "An improved framework for confound regression and filtering for control of motion artifact in the preprocessing of resting-state functional connectivity data.” *NeuroImage* 64 (1): 240–56. https://doi.org/10.1016/j.neuroimage.2012.08.052.

Tustison, N. J., B. B. Avants, P. A. Cook, Y. Zheng, A. Egan, P. A. Yushkevich, and J. C. Gee. 2010. “N4ITK: Improved N3 Bias Correction.” *IEEE Transactions on Medical Imaging* 29 (6): 1310–20. https://doi.org/10.1109/TMI.2010.2046908.

Zhang, Y., M. Brady, and S. Smith. 2001. “Segmentation of Brain MR Images Through a Hidden Markov Random Field Model and the Expectation-Maximization Algorithm.” *IEEE Transactions on Medical Imaging* 20 (1): 45–57. https://doi.org/10.1109/42.906424.

### Supplemental Tables

Table S1 *Imaging parameters*

| **Site** | **1** | **2** | **3** | **4** | **5** |
|---|---|---|---|---|---|
| Scanner | Philips Achieva 3.0T | Philips Ingenia 3.0T CX | GE 3.0T Discovery MR750 | Siemens MAGNETOM Skyra 3.0T | GE SIGNA 3.0T Premier |
| Head coil | 32-channel | 32-channel | 32-channel | 32-channel | 48-channel |
| **3D sagittal T1-weighted MPRAGE** | | | | | |
| TR (ms) a | 6.5 | 6.5 | 6.9 | 2300 | 2235 |
| TI (ms) | 900 | 900 | 900 | 900 | 900 |
| TE (ms) | 2.9 | 2.9 | 3 | 2 | 2.8 |
| Flip angle (⁰) | 9 | 9 | 9 | 9 | 9 |
| Voxel size (mm) | 1 x 1 x 1 | 1 x 1 x 1 | 1 x 1 x 1 | 1 x 1 x 1 | 1 x 1 x 1 |
| Matrix | 256 x 256 | 256 x 256 | 256 x 256 | 256 x 256 | 256 x 256 |
| **Resting-state fMRI** | | | | | |
| TR (ms) | 2200 | 2200 | 2200 | 2200 | 2200 |
| TE (ms) | 28 | 28 | 28 | 28 | 28 |
| # Slices | 44 | 44 | 42 | 42 | 42 |
| # Volumes | 275 | 275 | 275 | 272 | 272 |
| Voxel size (mm) | 3.3 × 3.3 × 3 | 3.3 × 3.3 × 3 | 3.3 × 3.3 × 3 | 3.3 × 3.3 × 3 | 3.3 × 3.3 × 3 |
| Slice gap (mm) | 0.3 | 0.3 | 0.3 | 0.3 | 0.3 |
| Matrix | 64 × 64 | 64 × 64 | 64 × 64 | 64 × 64 | 64 × 64 |
| Flip angle (⁰) | 80 | 80 | 80 | 80 | 80 |

Footnote: a) values for TR are highly variable due to different definitions of TR for this pulse sequence. Abbreviations: TR=repetition time, TE=echo time, TI=inversion time, T1w=T1 weighted, MPRAGE=magnetization-prepared rapid acquisition gradient-echo, fMRI=functional magnetic resonance imaging.

Table S2 *Demographic and clinical characteristics in early- vs late-onset OCD*

| Characteristic | OCD onset under 18 (N = 148) | OCD onset after 18 (N = 114) | p-value |
|---|---|---|---|
| Sex (N (%)) | | | 0.2* |
| Female | 75 (51%) | 68 (60%) | |
| Male | 73 (49%) | 46 (40%) | |
| Age (years) | 28.20 (7.85) | 31.28 (7.78) | 0.002† |
| Education (years) | 15.14 (2.91) | 15.32 (2.60) | 0.6† |
| IQ | 106.80 (11.94) | 101.89 (12.27) | 0.001† |
| Mean FD (mm) | 0.12 (0.06) | 0.12 (0.06) | 0.5† |
| Age of OCD onset (years) | 12.50 (3.31) | 23.36 (5.62) | <0.001† |
| Y-BOCS (M (SD)) | 24.65 (4.55) | 24.84 (5.30) | 0.8† |
| HAM-A (M SD)) | 12.76 (8.44) | 11.20 (7.26) | 0.11† |
| HAM-D (M (SD)) | 9.01 (5.86) | 8.44 (6.22) | 0.4† |
| Current comorbid anxiety disorder (N (%)) | 72 (49%) | 40 (35%) | 0.032* |
| Current comorbid MDD (N (%)) | 35 (24%) | 22 (19%) | 0.5* |
| SSRI/SNRI naïve (N (%)) | 79 (53%) | 68 (60%) | 0.3* |
| Benzodiazepines naïve (N (%)) | 132 (89%) | 104 (91%) | 0.7* |
| Antipsychotics naïve (N (%)) | 131 (89%) | 110 (96%) | 0.021* |
| Mood stabilizer naïve (N (%)) | 142 (96%) | 113 (99%) | 0.14* |
| CBT naïve (N (%)) | 104 (70%) | 94 (82%) | 0.029* |
| DY-BOCS (M (SD)) | | | |
| Harm & Aggression | 5.31 (4.74) – 1 missing | 5.22 (4.61) | 0.9† |
| Sexual & Religious | 4.51 (4.84) – 2 missing | 4.39 (5.04) | 0.9† |
| Symmetry & Ordering | 6.21 (4.42) – 1 missing | 5.21 (4.33) | 0.068† |
| Contamination | 6.40 (4.84) – 2 missing | 6.39 (5.13) | >0.9† |

*=Fisher's exact test; †=two-sample t-test. All p-values are uncorrected for multiple comparisons. Abbreviations: CBT=Cognitive Behavioral Therapy; DY-BOCS=Dimensional Yale-Brown Obsessive-Compulsive Scale; FD=Framewise Displacement; HAM-A=Hamilton Anxiety Rating Scale; HAM-D=Hamilton Depression Rating Scale; HC=Healthy Controls; IQ=Intelligence Quotient; M=Mean; MDD=Major Depressive Disorder; OCD=Obsessive-Compulsive Disorder; SD=Standard deviation; SNRI=Serotonin–Norepinephrine Reuptake Inhibitor; SSRI=Selective Serotonin Reuptake Inhibitor; Y-BOCS=Yale-Brown Obsessive Compulsive Scale.

Table S3 *Demographic and clinical characteristics in SSRI/SNRI naïve vs previously SSRI/SNRI medicated OCD*

| Characteristic | Previous SSRI/SNRI (N = 115) | SSRI/SNRI naive (N = 148) | p-value |
|---|---|---|---|
| Sex (N (%)) | | | >0.9* |

| | | | |
|---|---|---|---|
| Female | 63 (55%) | 81 (55%) | |
| Male | 52 (45%) | 67 (45%) | |
| Age (years) | 30.65 (7.93) | 28.68 (7.87) | 0.046† |
| Education (years) | 15.13 (2.92) | 15.28 (2.67) | 0.7† |
| IQ | 103.91 (12.42) | 105.28 (12.19) | 0.4† |
| Mean FD (mm) | 0.12 (0.06) | 0.13 (0.06) | 0.2† |
| Age of OCD onset (years) | 17.17 (6.76) | 17.27 (7.20) – 1 missing | >0.9* |
| OCD onset (N (%)) | | | 0.3* |
| 18- | 69 (60%) | 79 (54%) | |
| 18+ | 46 (40%) | 68 (46%) | |
| Y-BOCS (M (SD)) | 25.50 (5.06) | 24.16 (4.66) | 0.027† |
| HAM-A (M SD)) | 13.55 (7.90) | 10.93 (7.84) | 0.008† |
| HAM-D (M (SD)) | 9.83 (6.45) | 7.93 (5.51) | 0.012† |
| Current comorbid anxiety disorder (N (%)) | 58 (50%) | 92 (62%) | 0.061* |
| Current comorbid MDD (N (%)) | 32 (28%) | 25 (17%) | 0.036* |
| Benzodiazepines naïve (N (%)) | 94 (82%) | 143 (97%) | <0.001 * |
| Antipsychotics naïve (N (%)) | 96 (83%) | 146 (99%) | <0.001 * |
| Mood stabilizer naïve (N (%)) | 110 (96%) | 146 (99%) | 0.2* |
| CBT naïve (N (%)) | 77 (67%) | 122 (82%) | 0.006* |
| DY-BOCS (M (SD)) | | | |
| Harm & Aggression | 5.56 (4.91) – 1 missing | 5.07 (4.48) | 0.4† |
| Sexual & Religious | 4.58 (5.19) – 2 missing | 4.38 (4.70) | 0.8† |
| Symmetry & Ordering | 6.25 (4.52) – 1 missing | 5.45 (4.30) | 0.15† |
| Contamination | 6.88 (5.17) – 2 missing | 6.01 (4.76) | 0.2† |

*=Fisher's exact test; †=two-sample t-test. All p-values are uncorrected for multiple comparisons. Abbreviations: CBT=Cognitive Behavioral Therapy; DY-BOCS=Dimensional Yale-Brown Obsessive-Compulsive Scale; FD=Framewise Displacement; HAM-A=Hamilton Anxiety Rating Scale; HAM-D=Hamilton Depression Rating Scale; HC=Healthy Controls; IQ=Intelligence Quotient; MDD=Major Depressive Disorder; OCD=Obsessive-Compulsive Disorder; SNRI=Serotonin–Norepinephrine Reuptake Inhibitor; SSRI=Selective Serotonin Reuptake Inhibitor; Y-BOCS=Yale-Brown Obsessive Compulsive Scale.

Table S4 *Demographic and clinical characteristics in comorbid anxiety disorder versus non-anxiety disorder OCD*

| **Characteristic** | **No Anxiety Disorder (N = 150)** | **Comorbid Anxiety Disorder (N = 113)** | p-value |
|---|---|---|---|
| Sex (N (%)) | | | 0.13* |
| Female | 76 (51%) | 68 (60%) | |
| Male | 74 (49%) | 45 (40%) | |
| Age (years) | 29.90 (7.98) | 29.07 (7.90) | 0.4† |
| Education (years) | 15.07 (2.48) | 15.41 (3.12) | 0.4† |
| IQ | 104.71 (12.28) | 104.64 (12.34) | >0.9† |
| Mean FD (mm) | 0.13 (0.06) | 0.12 (0.06) | 0.5† |

| | | | |
|---|---|---|---|
| Age of OCD onset (years) | 18.18 (6.86) | 15.95 (7.01) – 1 missing | 0.011† |
| OCD onset (N (%)) | | | 0.032* |
| 18- | 76 (51%) | 72 (64%) | |
| 18+ | 74 (49%) | 40 (36%) | |
| Y-BOCS (M (SD)) | 24.79 (4.96) | 24.68 (4.78) | 0.9† |
| HAM-A (M SD)) | 10.51 (7.07) | 14.15 (8.60) | <0.001 † |
| HAM-D (M (SD)) | 8.02 (5.26) | 9.75 (6.76) | 0.025† |
| Current comorbid MDD (N (%)) | 21 (14%) | 36 (32%) | <0.001 * |
| SSRI/SNRI naïve (N (%)) | 92 (61%) | 56 (50%) | 0.061 |
| Benzodiazepines naïve (N (%)) | 135 (90%) | 102 (90%) | >0.9* |
| Antipsychotics naïve (N (%)) | 141 (94%) | 101 (89%) | 0.2* |
| Mood stabilizer naïve (N (%)) | 147 (98%) | 109 (96%) | 0.5* |
| CBT naïve (N (%)) | 110 (73%) | 89 (79%) | 0.4* |
| DY-BOCS | | | |
| Harm & Aggression | 4.81 (4.62) – 1 missing | 5.90 (4.68) | 0.062† |
| Sexual & Religious | 4.34 (4.91) – 1 missing | 4.63 (4.94) – 1 missing | 0.6† |
| Symmetry & Ordering | 4.77 (4.22) – 1 missing | 7.15 (4.30) | <0.001 † |
| Contamination | 6.10 (5.12) – 1 missing | 6.77 (4.72) – 1 missing | 0.3† |

*=Fisher's exact test; †=two-sample t-test. All p-values are uncorrected for multiple comparisons. Abbreviations: CBT=Cognitive Behavioral Therapy; DY-BOCS=Dimensional Yale-Brown Obsessive-Compulsive Scale; FD=Framewise Displacement; HAM-A=Hamilton Anxiety Rating Scale; HAM-D=Hamilton Depression Rating Scale; HC=Healthy Controls; IQ=Intelligence Quotient; M=Mean; MDD=Major Depressive Disorder; OCD=Obsessive-Compulsive Disorder; SD=Standard deviation; SNRI=Serotonin–Norepinephrine Reuptake Inhibitor; SSRI=Selective Serotonin Reuptake Inhibitor; Y-BOCS=Yale-Brown Obsessive Compulsive Scale.

Table S5 *Demographic and clinical characteristics in comorbid MDD versus non-MDD OCD*

| **Characteristic** | **No MDD (N = 206)** | **Comorbid MDD (N = 57)** | **p-value** |
|---|---|---|---|
| Sex (N (%)) | | | 0.001* |
| Female | 102 (50%) | 42 (74%) | |
| Male | 104 (50%) | 15 (26%) | |
| Age (years) | 29.78 (8.08) | 28.70 (7.41) | 0.3† |
| Education (years) | 15.29 (2.84) | 14.96 (2.51) | 0.4† |
| IQ | 105.17 (12.03) | 102.89 (13.12) | 0.2† |
| Mean FD (mm) | 0.12 (0.06) | 0.12 (0.07) | >0.9† |
| Age of OCD onset (years) | 17.30 (6.70) – 1 missing | 16.96 (8.05) | 0.8* |
| OCD onset (N (%)) | | | 0.5* |
| 18- | 113 (55%) | 35 (61%) | |
| 18+ | 92 (45%) | 22 (39%) | |

| | | | |
|---|---|---|---|
| Y-BOCS (M (SD)) | 24.16 (4.85) | 26.86 (4.39) | <0.001 * |
| HAM-A (M SD)) | 10.11 (6.56) | 19.18 (8.56) | <0.001 * |
| HAM-D (M (SD)) | 7.18 (4.85) | 14.49 (6.31) | <0.001 * |
| Current comorbid anxiety disorder (N (%)) | 77 (37%) | 36 (63%) | <0.001 * |
| SSRI/SNRI naïve (N (%)) | 123 (60%) | 25 (44%) | 0.036* |
| Benzodiazepines naïve (N (%)) | 188 (91%) | 49 (86%) | 0.3* |
| Antipsychotics naïve (N (%)) | 190 (92%) | 52 (91%) | 0.8* |
| Mood stabilizer naïve (N (%)) | 202 (98%) | 54 (95%) | 0.2* |
| CBT naïve (N (%)) | 157 (76%) | 42 (74%) | 0.7* |
| DY-BOCS | | | |
| Harm & Aggression | 4.94 (4.46) – 1 missing | 6.51 (5.21) | 0.042† |
| Sexual & Religious | 4.29 (4.87) – 2 missing | 5.07 (5.05) | 0.3† |
| Symmetry & Ordering | 5.54 (4.31) – 1 missing | 6.70 (4.66) | 0.095† |
| Contamination | 6.11 (4.85) – 2 missing | 7.39 (5.22) | 0.10† |

*=Fisher's exact test; †=two-sample t-test. All p-values are uncorrected for multiple comparisons. Abbreviations: CBT=Cognitive Behavioral Therapy; DY-BOCS=Dimensional Yale-Brown Obsessive-Compulsive Scale; FD=Framewise Displacement; HAM-A=Hamilton Anxiety Rating Scale; HAM-D=Hamilton Depression Rating Scale; HC=Healthy Controls; IQ=Intelligence Quotient; M=Mean; MDD=Major Depressive Disorder; OCD=Obsessive-Compulsive Disorder; SD=Standard deviation; SNRI=Serotonin–Norepinephrine Reuptake Inhibitor; SSRI=Selective Serotonin Reuptake Inhibitor; Y-BOCS=Yale-Brown Obsessive Compulsive Scale.
